# Role of intermediate care unit admission and non-invasive respiratory support during the COVID-19 pandemic: a retrospective cohort study

**DOI:** 10.1101/2020.07.17.20155929

**Authors:** Olivier Grosgurin, Antonio Leidi, Pauline Darbellay Farhoumand, Sebastian Carballo, Dan Adler, Jean-Luc Reny, Bernardo Bollen Pinto, Anne Rossel, Jacques Serratrice, Thomas Agoritsas, Jérôme Stirnemann, Christophe Marti

## Abstract

**Background:** The COVID-19 pandemic has led to shortage of Intensive Care Unit (ICU) capacity. We developed a triage strategy including non-invasive respiratory support and admission to the intermediate care unit (IMCU). ICU admission was restricted to patients requiring invasive ventilation.

**Objectives:** The aim of this study is to describe the characteristics and outcomes of patients admitted to the intermediate care unit.

**Method:** Retrospective cohort including consecutive patients admitted between March 28^th^ and April 27^th^ 2020. The primary outcome was the proportion of patients with severe hypoxemic respiratory failure avoiding ICU admission. Secondary outcomes included the rate of emergency intubation, 28-days mortality and predictors of ICU admission.

**Results:** One hundred fifty seven patients with COVID-19 associated pneumonia were admitted to the IMCU. Among the 85 patients admitted for worsening respiratory failure, 52/85 (61%) avoided ICU admission. In multivariate analysis, PaO2/FiO2 (OR 0.98; 95% CI 0.96 to 0.99) and Body Mass Index (OR 0.88; 95% CI 0.78 to 0.98) were significantly associated with ICU admission. No death or emergency intubation occurred in the intermediate care unit. Among the 72 patients transferred from the ICU, 60/72 (83%) presented neurological complications.

**Conclusions:** Non-invasive respiratory support including High-Flow Nasal Oxygen and continuous positive airway pressure prevents ICU admission for a large proportion of patients with COVID-19 hypoxemic respiratory failure. In the context of the COVID pandemic, intermediate care units may play an important role in preserving ICU capacity by avoiding ICU admission for patients with worsening respiratory failure and allowing early discharge of ICU patients.

## INTRODUCTION

The widespread expansion of the coronavirus disease 2019 (COVID-19) has led to a global pandemic and acute rise in hospital and Intensive Care Unit (ICU) admissions [1]. Many countries experienced capacity shortage in acute care and ICU, due to the sharp increase in admission of patients with acute hypoxemic respiratory failure (AHRF) and the prolonged length of ICU stay for patients requiring invasive mechanical ventilation [2-4]. At the peak of the pandemic, pessimistic projections predicted more than 3 million intensive care unit (ICU) admissions in the USA and more than 30 potential critically ill patients per ventilator place [3, 5]. Alarmingly poor outcomes were reported from Wuhan among patients requiring invasive mechanical ventilation, with mortality rates ranging between 81% and 97%[6]. The COVID-19 pandemic compelled health authorities of several countries to establish triage committees and restrictive admission criteria for ICU resource allocation [7, 5].

Intermediate care units (IMCUs), also known as high-dependency units, typically stand between regular ward and ICU. In Switzerland, these units provide continuous monitoring and an increased nurse to patient ratio compared to standard wards [8]. These units are able to provide several modalities of non-invasive respiratory support or vasopressors while patients requiring invasive mechanical ventilation are admitted to the ICU.

In the context of COVID-19 respiratory failure, IMCU may be the ideal structure to evaluate patient’s responsiveness to non-invasive respiratory support such as continuous positive airway pressure (CPAP) and high-flow nasal oxygen (HFNO) and to restrict ICU admission for those who fail to respond to non-invasive treatment. In addition, early discharge to the IMCU of ICU patients still requiring monitoring, heavy burden of care or specific procedures such as ventilation or tracheotomy weaning may contribute to preserve ICU capacity [9].

Our objective was to evaluate the contribution of IMCUs in the management of the COVID-19 respiratory pandemic. The primary objective was to evaluate the proportion of patients with severe hypoxemic respiratory failure admitted to the IMCU avoiding ICU admission and invasive mechanical ventilation.

Our secondary objectives were to assess the safety of this strategy by recording unexpected emergency intubation outside the ICU, mortality at 28 days, and the occurrence of sick-leaves among IMCU healthcare workers due to COVID-19 infections. We also assessed the role of transfer to the IMCU as a step-down unit after ICU stay, in order to preserve ICU capacity.

## METHODS

The study protocol was written according the STROBE Statement Checklist and is available on request (https://www.equator-network.org/). This observational study was approved by the ethics committee of our institution (CCER 2020-01087).

### Study Design and setting

We performed a retrospective cohort study of patients admitted to the IMCU of Geneva University Hospitals (HUG), Switzerland, during the COVID-19 pandemic. Consecutive patients admitted between March 28^th^ and April 27^th^ 2020 were included. In order to absorb the high volume of admissions, our IMCU capacity was progressively raised from 9 to 42 beds for patients with COVID-19 during the observation period. The capacity of the medical IMCU was increased by first transforming our coronary care and stroke units into medical IMCUs. Then, a new floor of IMC including 30 beds was established in collaboration with the Division of anaesthesiology. Additional Hamilton® T1 ventilators were provided by the Swiss Army forces. Nurses from the divisions of internal medicine and anaesthesiology, were attributed to this extended unit, as well as staff available after the stopping of surgical activity during the pandemic. The nurse-to-patient ratio was 2 to 5 with a supervising nurse (IMCU or anaesthesiology trained nurse) for every 10 patients. A dedicated respiratory therapist was available for the IMCU around the clock.

### Participants

Inclusion criteria

- COVID-19 pulmonary disease confirmed by SARS-Cov-2 reverse transcriptase polymerase chain reaction (rt-PCR) performed on either nasopharyngeal or oropharygeal swabs and image of pulmonary infiltrate AND Admission to IMCUs between March the 28th and April the 27th of 2020. Exclusion criteria (OR criteria)
- IMCU Admission for reasons other than COVID-19 pulmonary disease. Patients for which admission to the ICU was no more considered after decision of treatment de-escalation were excluded from the univariate and multivariate models exploring the likelihood of ICU admission.

### Procedures

In our institution, admission criteria to the IMCU were defined as the need for an inspired fraction of oxygen (FiO2) superior to 50% to maintain an oxygen saturation superior to 90% and the absence of signs of respiratory distress (defined as use of accessory muscles, intercostal retraction or alteration of mental status). Admission criteria to the IMCU and ICU during the study period are provided in table 1.

**Table 1.**
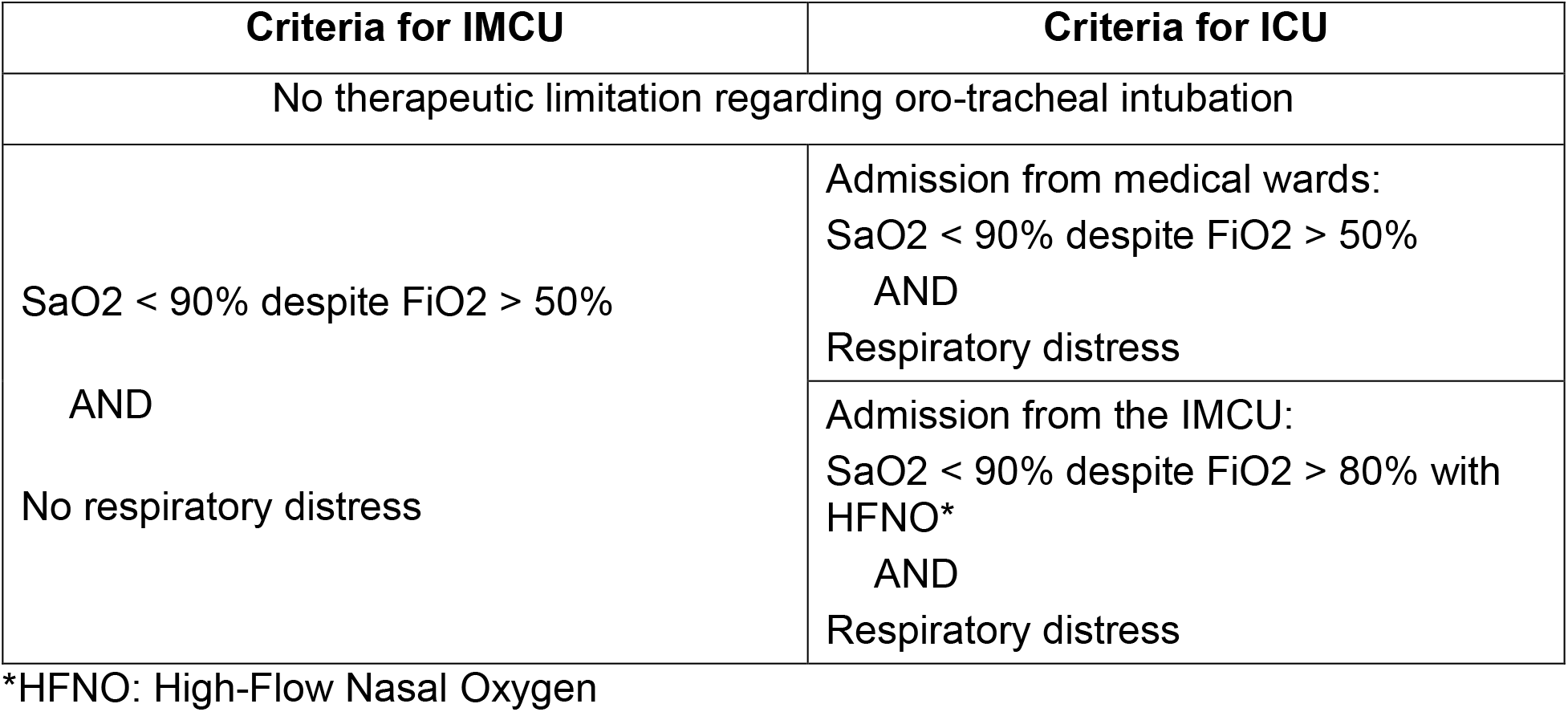
IMCU and ICU admission criteria at the University Hospital of Geneva during the COVID-19 pandemic.

HFNO was delivered using the Optiflow® system and Hamilton® T1 ventilators with humidifier. Oxygen Flow was set to 30 l/min, and higher flows (50 l/min) were allowed for patients showing improvement with continuous positive airway pressure (CPAP) but unable to tolerate it. FiO2 was titrated according to oxygen saturation aiming a saturation comprised between 90 and 94%.

CPAP (Continuous Positive Airway Pressure) was delivered during 6 sessions lasting for two hours through facemasks aiming for a positive end-expiratory pressure value of 10 cmH2O alternating with HFNO. An expiratory filter was systematically used. Using awake prone position was left at the discretion of the treating physician, and repeated according to tolerance and improvement in oxygenation parameters.

### Variables

Demographic (age, gender) variables, co-morbidities, Body mass index (BMI), respiratory parameters (FiO2, PaO2/FiO2, Respiratory rate and PaCO2 at admission), heart rate, symptoms duration, IMCU Length of stay (LOS) and receipt of antiviral therapy were extracted from the medical reports. Living status at 28 days was established based on medical records, administrative database of the Geneva province or contact with patients or relatives. The total number of cases hospitalized in our institution was extracted from our administrative database. The need for tracheotomy, ventilation and neurological complications for step-down patients was obtained from the medical records by study investigators.

### Data sources

The main data source was the electronic patient data system (DPI®) of the Geneva University Hospital. Data regarding the total number of patients admitted to the hospital and patients’ trajectories from the ward or emergency department to the ICU were obtained from the institutional administrative database. Data regarding the prevalence of absences due to COVID- 19 infection among IMCU health workers was obtained from our administrative database and included nurses and auxiliaries affiliated to these units during the study period.

### Outcomes

The primary outcome was the proportion of patients admitted to the ICU among patients admitted to the IMCU for worsening hypoxemia (i.e. patients admitted in a step-up strategy). Secondary outcomes included 28-days mortality, the number of death or emergent intubation in the IMCU and the rate of neurological complications and ICU readmissions among patients admitted to the IMCU from the ICU. Length of IMCU stay after admission from the ward and ICU readmission for patients discharged from the ICU were also recorded. The number of absences due to COVID-19 infections among healthcare workers was obtained based on an administrative database of our human resource department (Vision RH ®), in which absences during the study period due to COVID-19 infection were documented with a new specific code. COVID-19 related absences occurring from March 28^th^ until May 10^th^ 2020 were included in order to account for a two weeks incubation period.

### Statistical methods

The collection of data was performed by a research nurse (T. Mann), a biologist (L. Bayer) and two investigators (AL and OG).

Our study had a convenient sample size and no sample calculation was performed. Patients admitted to the IMCU of Geneva Hospitals since March 27^th^ were included as IMCU admission criteria for COVID-19 were established at this time. Recruitment was stopped on April 28^Th^ because the rate of admission of COVID-19 patients declined in our institution during the last week of this month and to allow us to complete 28-day follow-up at the end of May 2020.

### Statistical analysis

Demographic and descriptive statistics are provided in means with standard deviation (SD) or medians with interquartile ranges (IQR). Categorical and continuous variables were measured and compared between subsets of patients using chi^2^ and student *t* tests as appropriate. P-values for between group differences are provided with a statistical significance level defined as p-value < 0.05. In order to identify predictors of ICU admission, association between dependent variables and the primary outcome was assessed using univariate and multivariate logistic regression. The multivariate model was built using five pre-defined clinical variables (age, BMI, respiratory rate, heart rate and PaO2/FiO2). Analyses were conducted with IBM SPSS Statistics software (version 25) except for the multivariate model, for which R statistical software (R foundation) was used.

## RESULTS

During the study period, 2607 COVID infections were diagnosed in the Geneva province, 671 COVID patients were admitted to our Hospital and 63 were admitted to the ICU. 157 patients were admitted to the IMCU with COVID-associated pneumonia including 85 patients with worsening hypoxemic respiratory failure (step-up admissions) and 72 patients discharged from the ICU to the IMCU (step-down admissions) (Fig.1). The characteristics of patients admitted to the IMCU for treatment intensification are provided in table 2. Sixty-four out of 85 patients (74%) were male with a mean age of 63.9 years (SD12.8). The mean Arterial partial pressure of oxygen/Fraction of inspired Oxygen ratio (PaO2/FiO2) at IMCU admission was 133.5 mmHg (SD 54.4) and mean BMI was 27.9 kg/m^2^ (SD 5.7). Nineteen patients (22.4%) were admitted from the Emergency Department and 66 (77.6%) from the ward.

**Table 2.** Patient characteristics of IMCU step-up admissions according to the need for subsequent ICU transfer.

|  | **All Patients**<br>**(n = 85)** | **ICU**<br>**Transfer**<br>**(n=33)** | **No ICU**<br>**Transfer**<br>**(n=52)** | **p-value** |
| --- | --- | --- | --- | --- |
| **Age, years**<br>**mean (SD)** | 63.9 (12.8) | 65.3 (12.7) | 62.9 (12.9) | 0.42 |
| **Co-morbidities, any,**<br>**n, (%)** | 53 (61.6) | 21 (63.6) | 31 (59.6) | 0.14 |
| **BMI, kg/m<sup>2</sup>**<br>**mean (SD)** | 27.9 (5.7) | 26.8 (5.4) | 28.6 (5.9) | 0.20 |
| **FIO<sub>2</sub>**<br>**mean (SD)** | 0.59 (0.18) | 0.67 (0.2) | 0.54 (0.2) | **0.001** |
| **PaO<sub>2</sub>/FIO<sub>2</sub>, mmHg**<br>**mean (SD)** | 133.5 (54.4) | 113.8 (40.9) | 149.9 (58.9) | **0.004** |
| **PaCO<sub>2</sub>, mmHg**<br>**mean (SD)** | 31.7 (10.4) | 34.2 (7.7) | 30.1 (11.2) | 0.60 |
| **Respiratory rate,**<br>**mean (SD)** | 32.9 (5.0) | 34.1 (4.3) | 30.7 (5.1) | **0.002** |
| **Heart rate**<br>**mean (SD)** | 90.7 (17.7) | 92.7 (18.0) | 89.5 (17.2) | 0.42 |
| **Antiviral therapy,**<br>**any, n (%)** | 71 (83.5) | 28 (84.8) | 43 (82.7) | 0.068 |
| **Symptom duration,**<br>**days mean (SD)** | 10.2 (7.1) | 9.5 (4.6) | 11.5 (7.2) | 0.15 |
| **IMCU\* LOS<sup>+</sup>, days,**<br>**mean (SD)** | 2.8 (2.9) | 1.2 (0.7) | 4.1 (2.9) | **0.001** |
| **28-day mortality,**<br>**n (%)** | 10 (11.8) | 5 (15.2) | 5 (9.6) | 0.60 |

\*IMCU: intermediate care unit, <sup>+</sup>Length of stay

**Figure 1.**
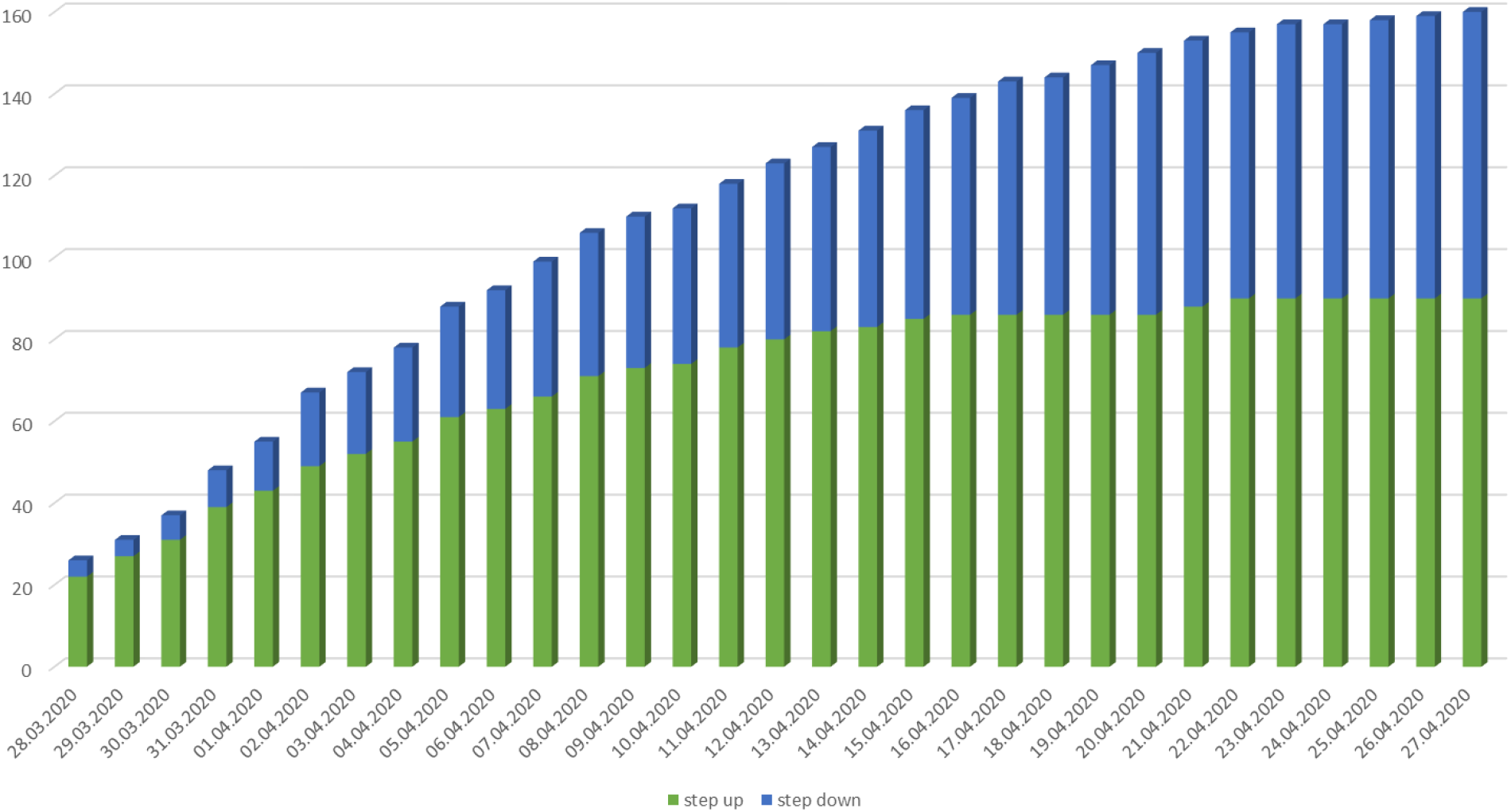
Cumulative number of “set up” and “set down” patients in the intermediate care unit

Of the 85 patients admitted for worsening hypoxemia (step-up admissions), 33 (39%) required ICU admission and 52 (61%) were discharged to the ward without being admitted to the ICU. Compared to patients discharged to the ward, patients requiring ICU admission required higher FiO2 at admission had lower PaO2/FiO2 ratios and higher respiratory rates (Table 2). The mean IMCU LOS was 1.2 days (SD 0.7) for patients requiring ICU admission and 4.1 days (SD 2.9) for those transferred to the ward. No unexpected death or emergent intubation occurred in the IMCU. The 28-day mortality was 15.2% (5/33) among patients requiring ICU admission and 9.6% (5/52) for patients transferred to the ward. The 5 IMCU patients who died without being admitted to the ICU died in the ward after decision of treatment de-escalation due to unfavourable course and discussion with the patient and/or relatives.

In univariate analysis, respiratory rate at IMCU admission (OR 1.16; 95% CI 1.06 to 1.31) and reduced PaO2/FiO2 ratio (OR 0.98; 95%CI 0.97 to 0.99) were significantly associated with ICU admission. In the multivariate model including age, BMI, heart rate, respiratory rate and PaO2/FiO2, PaO2/FiO2 (OR 0.98, 95%CI 0.96 to 0.99) and BMI (OR 0.88; 95% CI 0.78 to 0.98) were significantly associated with ICU admission (Table 3).

**Table 3:** Association between patient characteristics and transfer to the ICU among IMCU step-up admissions.

| **Variable** | **Univariate OR**<br>**(95%CI)** | **Multivariate OR** |
| --- | --- | --- |
| Age | 1.01 (0.98 to 1.05) | 1.03 (0.99 to 1.09) |
| BMI* (kg/m<sup>2</sup>) | 0.94 (0.85 to 1.02) | 0.88 (0.78 to 0.98) |
| Heart rate (min<sup>-1</sup>) | 1.01 (0.99 to 1.04) | 1.00 (0.97 to 1.03) |
| Respiratory rate | 1.16 (1.06 to 1.31) | 1.07 (0.95 to 1.23) |
| PaO<sub>2</sub>/FiO<sub>2</sub><br>(mmHg) | 0.98 (0.97 to 0.99) | 0.98 (0.96 to 0.99) |

\*BMI: Body mass index

Seventy-two patients were admitted to the IMCU after discharge from the ICU. Characteristics of these patients are provided in table 4. These patients were predominantly males and 52/72 (72.2 %) had at least one documented pre-existing condition. The mean ICU LOS among these step-down patients was 16.1 days (SD 7.6) and 70/72 (97.2%) received invasive mechanical ventilation during the ICU stay. A large majority of these step-down patients presented at least one neurological complication (Table 4). ICU readmission occurred in 5 (6.9%) patients and 28- day mortality in none.

**Table 4.** Characteristics of IMCU step-down patients.

|  |  |
| --- | --- |
| **Total number of patients (%)** | **72 (100)** |
| Mean Age, years (SD) | 62.8 (12.2) |
| Male gender, n (%) | 60 (83.3) |
| Co-morbid conditions, any, n (%) | 52 (72.2) |
| • Cancer | 0 (0) |
| • Hypertension | 43 (59.7) |
| • Cardiac disease | 18 (25.0) |
| • Diabetes | 21 (29.2) |
| • COPD | 3 (4.2) |
| • Renal failure | 4 (5.6) |
| ICU Length of stay, n (SD) | 16.1 (7.6) |
| Invasive mechanical ventilation in the ICU, n (%) | 70 (97.2) |
| Tracheotomy | 13 (18.1) |
| • Also requiring invasive mechanical ventilation at IMCU* admission | 8 (11.1) |
| Neurological complication, any, n (%) | 60 (83.3) |
| • Delirium or confusion | 44 (61.1) |
| • Polyneuropathy | 45 (62.5) |
| • Stroke | 7 (9.7) |
| Venous thrombo-embolism, n (%) | 21 (29.2) |
| ICU readmission, n (%) | 5 (6.9) |
| 28-day Mortality n (%) | 0 (0) |

\*IMCU: intermediate care unit

From March 28^th^ to May 15^th^, 5 out of 103 (4.9%) healthcare workers dedicated to the IMCU had a COVID-19 related absence in the administrative database.

## DISCUSSION

Our study suggests that non-invasive respiratory support including HFNO and CPAP delivered in the IMCU may contribute to prevent ICU admission and invasive mechanical ventilation in a majority of patients with COVID-19-associated AHRF without signs of respiratory distress.

Despite severe hypoxemia (mean PaO2/FiO2 133.5 mmHg), about 60% of patients in our cohort were transferred back to the ward without the need for an ICU admission. This finding may be highly relevant in the context of a respiratory pandemic where ICU capacity is a critical issue.

The role of non-invasive ventilation in AHRF has been debated over the last decades as delaying intubation for patients requiring invasive mechanical ventilation has been reported to be associated with negative patient outcome [10]. Non-invasive ventilation with inspiratory aid is usually not recommended, because of the risk of self-inflicted volume traumatism in patients with spontaneous breathing [11, 12]. However, HFNO has been reported to reduce 90-day mortality in a randomised control trial including 310 patients with AHRF compared to standard oxygen or non-invasive ventilation [13]. In a recent network meta-analysis including 25 studies and 3804 patients, both non-invasive ventilation and HFNO reduced the rate of oro-tracheal intubation among patients with AHRF [14]. The network effect estimates suggested that helmet non-invasive ventilation might be the most effective option through allowing higher levels of positive end-expiratory pressure and reduced inspiratory efforts [15, 16]. However, this evidence relies mainly on indirect comparisons and on studies including a large proportion of patients with community acquired pneumonia or immunosuppression. These patients may differ from COVID-19 associated AHRF. Indeed, different phenotypes of acute lung injury have been reported among patients with COVID-19 associated AHRF including near preserved lung compliance and low ventilation-to-perfusion ratios [17].

In our institution, we decided to combine HFNO using nasal cannulas and CPAP delivered through a facial mask for pragmatic reasons. These two modalities were possible using the same ventilator (Hamilton® T1) and our IMCU nurses and respiratory therapists are more used to this interface. Because some concern has been raised regarding the risk of potential aerosolisation and health-care workers contamination during HFNO or non-invasive ventilation, we aimed to reduce this risk by using personal protective equipment (PPE) including glasses, FFP2 masks, gloves and protective overdresses in the entire zone of care. Moreover, oxygen flow was restricted to 30 l/min using HFNO, except for patients with significant improvement during CPAP but unable to tolerate the mask interface. Using these precautions, the rate of contamination among IMCU healthcare workers in our study was low and similar to the rate of infection in the Geneva province according to a large seroprevalence study conducted at our institution [18]. However, our study only considered symptomatic infections leading to the absence of collaborators and could therefore fail to identify asymptomatic infections. Moreover, the potential source of contamination was not investigated.

Other emerging evidence regarding the role of non-invasive respiratory support in the context of COVID-19 AHRF has recently been reported. In a retrospective study comparing two consecutive periods, Oranger et al. reported a rate reduction of invasive mechanical ventilation in the intervention period using CPAP compared to the control period using increasing supplemental oxygen [19]. Similarly, HFNO or CPAP combined with patient self-proning have been reported to improve oxygenation in several case series [20-22].

In our cohort, combining HFNO, CPAP and patient self-proning in an IMCU resulted in a high proportion of patients avoiding invasive mechanical ventilation in the ICU. A key consideration is the use of standardized criteria for IMCU admission based on the absence of signs of respiratory distress. This strategy is in accordance with the World Health Organisation (WHO) guidance for the use of HFNO in COVID-19 associated AHRF recommending against the use of HFNO for patients with hypercapnia, haemodynamic instability, multi-organ failure or abnormal mental status [23].

As previously reported by others, overweight was highly prevalent among patients developing AHRF requiring admission to our IMCU [24]. Unexpectedly, increasing BMI was inversely associated with ICU admission in our cohort. This apparent paradox may suggest that obese patients might respond more favourably to non-invasive respiratory support than patients with other mechanisms of AHRF.

During the study period, about one half of patients admitted to the ICU were admitted from the IMCU, the remaining being admitted from the ED (21/63) or directly from the ward (9/63) suggesting that these criteria may be applied to an important proportion of patients. These empirically established criteria were based on the observation that a subset of patients with COVID-19 AHRF showed an excellent tolerance to hypoxemia (sometimes referred to as “happy hypoxemia”).

Finally, avoiding invasive mechanical ventilation in the context of the COVID-19 pandemic does not only preserve ICU capacity, but may also benefit patients by preventing invasive mechanical ventilation-related complications. In particular, the prevalence of neurological complications was high among ICU survivors discharged to the IMCU compared to IMCU patients transferred to the ward. This difference may be partly explained by the selection of patients with increased COVID-19 disease severity to be transferred to the ICU and the fact that the occurrence of obtundation or altered mental status may have prompted ICU admission in a subset of patients. However the high prevalence of muscle weakness, ICU acquired polyneuropathy and delirium among ICU survivors may also result of the need for prolonged sedation, use of paralysing agents and invasive mechanical ventilation.

Our study has several limitations. First, we used a convenient sample size, limiting the number of event and our ability to exhaustively identify predictors of ICU admission. Second, IMCUs do not exist worldwide, precluding the generalizability of our observation. However, we believe that our experience could be an interesting model in other settings, and may apply to similar structures such as respiratory care or high-dependency units. Third, we used a combination of HFNO, CPAP and self-prone position. The duration of interventions was not systematically recorded, preventing us to analyse the benefit of each modality individually. Finally, our observational design precludes drawing definite conclusion about the beneficial effect of IMCU admission or non-invasive ventilation in the absence of a control group. Further research, including interventional controlled studies, are needed to confirm our findings.

## CONCLUSION

Our IMCU admitted patients with severe COVID-19 presenting an AHRF yet without respiratory distress. A treatment combining HFNO, CPAP and self-prone position prevented a large proportion of patients from being transferred to the ICU. This model of care may be beneficial to preserve ICU capacity and reduce the complications associated with invasive mechanical ventilation.

## Data Availability

Data is available in our secured computerized database.

## ACKNOWLEDGEMENT

We thank our study nurses, Tamara Mann and Laurence Bayer, for data collection, and Prof Bernhard Walder for his advice and clinical collaboration. We would like to thank sincerely all nurses, residents and other health care providers who allowed us to manage the COVID pandemic at the IMCU of the Geneva University Hospital, as well as the heads of the Departments of Medicine and Acute Care for their trust and support during this challenging period.

## STATEMENT OF ETHICS

This study was conducted ethically in accordance with the World Medical Association Declaration of Helsinki. It was approved by the ethics committee of our institution (CCER 2020- 01087).

## CONFLICT OF INTEREST STATEMENT

The authors have no conflicts of interest to declare.

## FUNDING SOURCE

This study had no financial support.

## AUTHOR CONTRIBUTIONS

OG, CM and AL participated to data acquisition, design and drafting of the manuscript. OG and JST performed the statistical analyses. All authors participated to study conception, data interpretation and critical revision of the manuscript for important intellectual content. All authors approved the version submitted for publication and agreed to be accountable regarding accuracy and integrity of any part of the work. CM and OG had full access to the data and take responsibility for the integrity and the accuracy of the data.

## Notes

### Competing Interest Statement

The authors have declared no competing interest.

### Funding Statement

The authors have no conflicts of interest to declare. This study had no financial support.

### Author Declarations

This observational study was approved by the ethics committee of our institution (CCER 2020-01087).

